# FRONTAL WHITE MATTER CHANGES INDICATE RECOVERY WITH INPATIENT TREATMENT IN HEROIN ADDICTION

**DOI:** 10.1101/2024.06.10.24308719

**Authors:** Pierre-Olivier Gaudreault, Sarah G King, Yuefeng Huang, Ahmet O. Ceceli, Greg Kronberg, Nelly Alia-Klein, Rita Z. Goldstein

**Author notes:** **Corresponding author:** Rita Z. Goldstein, PhD Mount Sinai Professor in Neuroimaging of Addiction Department of Psychiatry (primary) and Department of Neuroscience (secondary) Chief, Neuropsychoimaging of Addiction and Related Conditions (NARC) Research Program Icahn School of Medicine Gustave L. Levy Place, Box 1230 New York, NY 10029.

## Abstract

**Importance:** Amidst an unprecedented opioid epidemic, identifying neurobiological correlates of change with medication-assisted treatment of heroin use disorder is imperative. Distributed white matter (WM) impairments in individuals with heroin use disorder (iHUD) have been associated with increased drug craving, a reliable predictor of treatment outcomes. However, little is known about the extent of whole-brain structural connectivity changes with inpatient treatment and abstinence in iHUD.

**Objective:** To assess WM microstructure and associations with drug craving changes with inpatient treatment in iHUD (effects of time/re-scan compared to controls; CTL).

**Design:** Longitudinal cohort study (12/2020-09/2022) where iHUD and CTL underwent baseline magnetic resonance imaging (MRI#1) and follow-up (MRI#2) scans, (mean interval of 13.9 weeks in all participants combined).

**Setting:** The iHUD and CTL were recruited from urban inpatient treatment facilities and surrounding communities, respectively.

**Participants:** Thirty-four iHUD (42.1yo; 7 women), 25 age-/sex-matched CTL (40.5yo; 9 women).

**Intervention:** Between scans, inpatient iHUD continued their medically-assisted treatment and related clinical interventions. CTL participants were scanned at similar time intervals.

**Main Outcomes and Measures:** Changes in white matter diffusion metrics [fractional anisotropy (FA), mean (MD), axial (AD), and radial diffusivities (RD)] in addition to baseline and cue-induced drug craving, and other clinical outcome variables (mood, sleep, affect, perceived stress, and therapy attendance).

**Results:** Main findings showed HUD-specific WM microstructure changes encompassing mostly frontal major callosal, projection, and association tracts, characterized by increased FA (.949<1- p<.986) and decreased MD (.949<1-p<.997) and RD (.949<1-p<.999). The increased FA (r=- 0.72, p<.00001) and decreased MD (r=0.69, p<.00001) and RD (r=0.67, p<.0001) in the genu and body of the corpus callosum and the left anterior corona radiata in iHUD were correlated with a reduction in baseline craving (.949<1-p<.999). No other WM correlations with outcome variables reached significance.

**Conclusions and Relevance:** Our findings suggest whole-brain normalization of structural connectivity with inpatient medically-assisted treatment in iHUD encompassing recovery in frontal WM pathways implicated in emotional regulation and top-down executive control. The association with decreases in baseline craving further supports the relevance of these WM markers to a major symptom in drug addiction, with implications for monitoring clinical outcomes.

**Key points:** *Question:* Does white matter (WM) microstructure change with medication-assisted treatment in individuals with heroin use disorder (iHUD)?

*Findings:* In this longitudinal cohort study, diffusion MRI was acquired in 34 inpatient iHUD and 25 healthy controls (CTL) twice, separated by a mean of 13.9 weeks. We found HUD-specific WM microstructure changes with time, characterized by increased anisotropy and decreased diffusivity in fronto-striatal WM pathways. These changes were correlated with decreased baseline drug craving with treatment.

*Meaning:* Frontal WM changes and associated drug craving decreases suggest brain-behavior recovery with inpatient treatment in iHUD, potentially contributing to reduced drug use and sustained abstinence.

## Introduction

North America is currently experiencing an opioid epidemic that caused approximately 107,000 fatal overdoses in 2021^1^, with opioid use disorders associated with particularly high relapse rates (e.g., 91%^2^ as compared to 40-60% in other substance use disorders)^3^. The complexity in treating heroin use disorder (HUD) stems in part from drug-related craving, the intense subjective urge to use drugs^4^, a reliable predictor of treatment outcomes (e.g., sustained abstinence and propensity to relapse)^5,6^. Identifying neurobiological correlates of change with treatment, including in craving reductions, is therefore a major need for individuals with HUD (iHUD).

Studies investigating neurobiological mechanisms underlying drug craving and other treatment outcomes show specific impairments in the cortico-striatal and mesolimbic pathways and neuronal projections reaching prefrontal regions^7,8^. However, these studies have mostly relied on MRI-based functional neuroimaging-derived task-based regional changes^9–11^ and whole-brain functional connectivity^12,13^, with less emphasis on structural connectivity underlying clinical symptoms. We recently reported widespread whole-brain WM impairments in individuals with cocaine and heroin use disorders as evidenced by decreased fractional anisotropy (FA), a measure of white matter organizational coherence, and increased mean (MD), axial (AD), and radial (RD) diffusivities, measures that have been associated with axonal damage and demyelination, compared to healthy control participants (CTL)^14^. Across all subjects, these WM deficits were associated with longer duration of regular use, suggestive of a cumulative and/or persistent effect, as well as with higher state-like (baseline) craving measures, highlighting the importance of current symptom severity^14^. While this latter study added to the growing literature showing whole-brain WM damage in iHUD^15–19^, far less research has been dedicated to longitudinally assessing these impairments and their potential for recovery with time into abstinence or treatment, as associated with parallel putative changes in substance use disorder symptoms including craving.

A recent systematic review of longitudinal structural (gray and white matter) and functional (including EEG, fMRI, and neurochemical) neuroimaging studies suggested recovery with abstinence across most substance use disorders, with the onset of structural recovery preceding neurochemical and functional recoveries^20^. Specifically in HUD, the earliest evidence for initial WM recovery was provided in a preliminary report of a no longer detectable decreased FA in the frontal and cingulate gyri when compared to CTL after one month of abstinence as compared to baseline (3 days of abstinence) in 20 individuals with HUD^21^. Recent atlas-based studies using group-averaged tractography masks reported increased FA in specific cortico-striatal WM tracts from approximately two to eight months of abstinence in two groups of iHUD, which was associated with decreased drug craving scores at follow-up^22^ and predicted longitudinal craving changes^23^. However, these studies focused on iHUD treated only with psychoeducational and social/physical rehabilitation, and therefore less is known about the effect of the US system of standard-of-care medication-assisted treatment (MAT) on WM microarchitecture.

The aim of this study was to longitudinally assess WM changes with inpatient MAT in iHUD, compared to matched CTL. Using a tensor-based voxelwise approach to evaluate whole-brain changes in WM, we expected normalization of diffusion metrics including increased FA and decreased MD, AD, and RD with greater time in treatment (≈14 weeks interval) in iHUD, and minimal changes in CTL over an equivalent interval. Since different craving measures provide distinct indicators of treatment outcomes^6^, we tested the association of WM changes with changes in both baseline/state-like (i.e., assessed without cue exposure) and cue-induced/dynamic craving. A secondary objective included testing associations between these WM changes and select behavioral clinical measures including mood, sleep, affect, perceived stress, and therapy attendance.

## Materials and Methods

### Participants

Thirty-four iHUD undergoing inpatient MAT (methadone N=30; buprenorphine N=4) were recruited from a substance use disorder rehabilitation facility (Samaritan Daytop Village, Queens, NY), where they attended courses on relapse prevention, “Seeking Safety” therapy, and anger management. From this sample, 21 iHUD were included in our recent cross-sectional report^14^. Twenty-five age- and sex-matched CTL were recruited through advertisements and word-of-mouth from the same communities. All participants were recruited as part of an ongoing clinical trial (NCT04112186) evaluating longitudinal therapy-specific neuroimaging changes in HUD (intervention-related outcomes to be reported after trial completion, see Supplementary Materials and Methods). See Table 1 for sample descriptive statistics. All subjects completed both baseline and follow-up assessments (mean time between-scans=13.9±5.8 weeks). All iHUD met DSM-5 criteria for opioid use disorder (with heroin as their primary drug of choice/reason for treatment) and were stabilized on MAT (confirmed via urine toxicology in both sessions) with a mean baseline abstinence of 176.6±215.3 days. Exclusion criteria are described in the Supplementary Materials and Methods, along with data on psychiatric comorbidities. All comorbidities in iHUD were either in partial or sustained remission, and CTL had no current or prior substance use disorder or other psychiatric conditions. All participants provided written informed consent; study procedures were approved by the institutional review board of the Icahn School of Medicine at Mount Sinai.

**Table 1.** Baseline demographic characteristics, neuropsychological measures, and drug use variables.

|  | Healthy Controls<br>CTL, N = 25 | Heroin use disorder<br>HUD, N = 34 | Statistical analyses |  |
| --- | --- | --- | --- | --- |
|  |  |  | Test | p value |
| **Demographics** |  |  |  |  |
| Age, mean (SD), years | 40.5 (11.0) | 42.1 (9.0) | $t_{(57)} = -0.6$ | $p = .542$ |
| Gender |  |  |  |  |
| Men, No. (%) | 16 (64) | 27 (79) |  |  |
| Women, No. (%) | 9 (36) | 7 (21) | $\chi^2 = 1.7$ | $p = .188$ |
| Education, mean (SD), years | 16.2 (3.3) | 12.2 (1.9) | $t_{(57)} = 5.8$ | **$p < .001$** |
| Race |  |  |  |  |
| Black or African American, No. (%) | 8 (32) | 3 (9) |  |  |
| White, No. (%) | 10 (40) | 17 (50) | $\chi^2 = 5.2$ | $p = .075$ |
| Other & Mixed, No. (%) | 7 (28) | 14 (41) |  |  |
| Days between scans, mean (SD) | 91.1 (50.3) | 101.7 (31.7) | $t_{(57)} = -1.0$ | $p = .328$ |
| Intracranial volume, mean (SD), liters | 1.62 (.13) | 1.70 (.15) | $t_{(57)} = -2.3$ | $p = .028$ |
| **Neuropsychological and self-reported tests** |  |  |  |  |
| WRAT – Reading scale standard score, mean (SD) | 108.8 (9.3) | 95.0 (12.1) | $t_{(56)} = 4.7$ | **$p < .001$** |
| WASI – Matrix Reasoning scaled score, mean (SD) | 12.0 (3.2) | 10.5 (3.1) | $t_{(56)} = 1.8$ | $p = .082$ |
| Handedness |  |  |  |  |
| Right, No. (%) | 18 (72) | 27 (79) |  |  |
| Left, No. (%) | 7 (28) | 7 (21) | $\chi^2 = 0.4$ | $p = .508$ |
| Baseline visit depression (BDI)<sup>a</sup>, mean (SD) | 3.3 (4.7) | 13.8 (11.1) | $t_{(57)} = -4.5$ | **$p < .001$** |
| Baseline visit anxiety (BAI)<sup>a</sup>, mean (SD) | 2.1 (4.1) | 10.6 (9.3) | $t_{(57)} = -4.3$ | **$p < .001$** |
| **Drug use variables** |  |  |  |  |
| Regular nicotine use, current, No. (%) | 1 (4) | 33 (97) | b | b |
| Fagerström test for nicotine dependence, mean (SD) | <sup>b</sup> | 3.6 (1.6) | n/a | n/a |
| Cigarettes use during past 30 days, mean (SD), days | <sup>b</sup> | 28.8 (5.4) | n/a | n/a |
| Cigarettes per day, No. (%)<sup>(10 or less/11 to 20/21 or more)</sup> | <sup>b</sup> | 28 (82) / 5 (15) / 1 (3) | n/a | n/a |
| Regular alcohol use, yes, No. (%) | 14 (56) | 21 (62) | $\chi^2 = 0.2$ | $p = .656$ |
| Lifetime alcohol regular use, mean (SD), years | 12.3 (11.2) | 14.5 (10.8) | $t_{(32)} = -0.6$ | $p = .580$ |
| Alcohol use during past 30 days, mean (SD), days | 6.9 (8.2) | 0.0 (0.0) | $t_{(33)} = 3.9$ | **$p < .001$** |
| Regular cannabis use, yes, No. (%) | 7 (28) | 21 (62) | $\chi^2 = 8.7$ | **$p = .003$** |
| Lifetime cannabis regular use, mean (SD), years | 3.0 (2.1) | 13.0 (7.9) | $t_{(26)} = -3.3$ | **$p < .001$** |
| Cannabis use during past 30 days, mean (SD), days | 0.4 (1.1) | 0.0 (0.0) | $t_{(26)} = 1.8$ | $p = .083$ |
| **Heroin use disorder** |  |  |  |  |
| Age of first use, mean (SD), years | n/a | 24.2 (8.4) | n/a | n/a |
| Years of regular use, mean (SD) | n/a | 10.7 (7.2) | n/a | n/a |
| Current abstinence, mean (SD), days | n/a | 176.6 (215.3) | n/a | n/a |
| Heroin use during past 30 days, mean (SD), days | n/a | 0.4 (1.2) | n/a | n/a |
| Self-reported Methadone dosage, mean (SD), mg | n/a | 107.2 (57.4)<sup>c</sup> | n/a | n/a |
| Self-reported Buprenorphine dosage, mean (SD), mg | n/a | 10.0 (9.7)<sup>d</sup> | n/a | n/a |
| Severity of Dependence, mean score (SD) | n/a | 10.9 (3.6) | n/a | n/a |
| Withdrawal symptoms, mean SOWS score (SD) | n/a | 3.7 (4.9) | n/a | n/a |
| Subjective craving, mean HCQ score (SD) | n/a | 39.9 (12.6) | n/a | n/a |
| **Treatment adherence** |  |  |  |  |
| Therapy attendance, mean (SD), hours | n/a | 10.6 (4.9) | n/a | n/a |

**Notes:** <sup>a</sup>The averaged baseline depression and anxiety symptoms are presented here. Time effects on these measures in individuals with HUD are presented in Table 2. <sup>b</sup>Since only one healthy control participant reported currently using nicotine, no statistical tests were done to compare the groups on measures of nicotine use. <sup>c</sup>Methadone, N=30, <sup>d</sup>Buprenorphine, N=4. WRAT: Wide Range Achievement Test; WASI: Wechsler Abbreviated Scale Intelligence; SOWS: Short Opiate Withdrawal Scale; HCQ: Heroin Craving Questionnaire. To correct for multiple comparisons, p values were considered significant at $p < 0.005$ (0.05/11 statistical tests) for the demographics/neuropsychological tests and at $p < 0.008$ for the drug use variables (0.05/6 tests). Missing data for each variable: 1 datapoint for WASI; 1 datapoint for WRAT; 1 datapoint for years of alcohol use; 3 datapoints for cannabis regular use; 2 datapoints for years of heroin regular use.

### Clinical Assessments

Highly trained clinical coordinators supervised by clinical psychologists conducted diagnostic evaluations that included the Mini International Neuropsychiatric Interview 7^th^ edition^24^ and the Addiction Severity Index^25^. Withdrawal symptoms were assessed with the Short Opiate Withdrawal Scale^26^ and self-reported subjective heroin craving with the Heroin Craving Questionnaire (a modified version of the Cocaine Craving Questionnaire)^27,28^. Dependence severity was assessed with the Severity of Dependence Scale^29^ and nicotine dependence with the Fagerström Test for Nicotine Dependence^30^. In addition to demographic characteristics and clinical drug use variables, Table 1 presents other neuropsychological and self-reported measures as well as the iHUD treatment adherence, measured with the number of hours of group-therapy attended. Table 2 shows behavioral measures at both imaging timepoints in iHUD (separated by 101.7±31.7 days, equivalent to a mean of 14.5 weeks) and reports between-scan comparisons in depression and anxiety symptoms, sleep quantity (hours) and sleep quality the night before the scan, positive and negative affect, and perceived stress (see Supplementary Materials and Methods). In addition to those measures, assessment of baseline drug craving was conducted at both timepoints at the start of a drug cue reactivity fMRI task (“Please rate how strong your desire for heroin is currently on a scale of 0-9”), reported elsewhere^9^. Following this baseline craving assessment, three stimulus ratings were measured after the MRI procedures to assess specific aspects of dynamic, cue-induced craving: picture-induced drug arousal (“*How emotional do you feel about the picture*”, a 5-point scale ranging from “calm, no emotion” to “extremely emotional”) and picture-induced drug craving (“*How strong is your desire to use the substance*”, a 5-point scale ranging from “no desire” to “extreme desire”) ratings of the same drug pictures used in the cue-reactivity fMRI task, and scene-induced drug craving intensity ratings in response to 34 3-sec clips of a naturalistic drug-related movie presented during an fMRI task^31^.

**Table 2.** Behavioral and drug-related craving measures repeated after ∼15-weeks of inpatient treatment in individuals with heroin use disorder.

|  | Heroin Use Disorder<br>(HUD, N = 34) |  | Statistical analyses |  |
| --- | --- | --- | --- | --- |
|  | MRI#1 | MRI#2 | Time effect | p value |
| ***Behavioral measures*** |  |  |  |  |
| Depression (BDI), mean score (SD) | 13.8 (11.1) | 12.0 (10.7) | $t_{(31)} = 1.7$ | $p = .095$ |
| Anxiety (BAI), mean score (SD) | 10.6 (9.3) | 10.7 (10.4) | $t_{(32)} = 0.1$ | $p = .911$ |
| Sleep quantity, mean (SD), hours | 6.9 (1.7) | 6.4 (1.9) | $t_{(33)} = 1.6$ | $p = .118$ |
| Sleep quality, mean score (SD) | 3.5 (1.2) | 3.7 (1.2) | $t_{(33)} = -0.5$ | $p = .649$ |
| PANAS – Positive, mean score (SD) | 31.3 (8.9) | 34.2 (8.6) | $t_{(32)} = -2.0$ | $p = .054^a$ |
| PANAS – Negative, mean score (SD) | 20.6 (8.9) | 17.9 (6.4) | $t_{(32)} = 1.9$ | $p = .066^a$ |
| PSS, mean score (SD) | 27.9 (7.3) | 26.9 (7.1) | $t_{(32)} = 0.8$ | $p = .443$ |
| ***Craving variables*** |  |  |  |  |
| ***Baseline (state-like) measure*** |  |  |  |  |
| Pre-task craving, mean rating (SD) | 3.3 (2.4) | 2.0 (2.7) | $t_{(33)} = 3.5$ | **$p = .001$** |
| ***Cue-Induced (dynamic) measures*** |  |  |  |  |
| Drug cues ratings – arousal, mean (SD) | 2.3 (.84) | 1.9 (.79) | $t_{(33)} = 2.5$ | $p = .019$ |
| Drug cues ratings – craving, mean (SD) | 1.9 (1.0) | 1.5 (.75) | $t_{(33)} = 2.4$ | $p = .023$ |
| Movie scene-induced craving intensity, mean (SD) | 1.3 (1.1) | .79 (.86) | $t_{(31)} = 3.9$ | **$p < .001$** |

**Notes:** Data expressed as frequencies or means ± standard deviation (SD). BDI: Beck Depression Inventory; BAI: Beck Anxiety Inventory; PANAS: Positive and Negative Affect Schedule; PSS: Perceived Stress Scale. To correct for multiple comparisons, p values significance was set at $p < 0.007$ (.05/7 statistical tests) for the behavioral measures and $p < 0.013$ (.05/4 statistical tests) for the drug craving variables. Comparisons reaching $p < .05$ were reported as trends. <sup>a</sup>Although independent longitudinal comparisons for positive and negative affect questionnaires only showed trends for increase and decrease between sessions, respectively, a time × affect ANOVA showed a significant interaction ( $F(1,32)=5.28$ , $p=.028$ ) with simple effects showing significant differences between both affect states at both time points (Positive>Negative, $p<.001$ ). Missing data for each variable: 2 data points for BDI (MRI#2); 1 data point for BAI (MRI#2); 1 data point PANAS (MRI#2); 1 data point for PSS (MRI#2); 2 data points for scene-induced craving intensity (1 for MRI#1 and 1 for MRI#2).

### MRI acquisition and Diffusion Tensor Imaging processing

MRI acquisitions were performed using a Siemens 3.0 Tesla Skyra scanner (Siemens Healthineers AG, Erlangen, Germany) with a 32-channel head coil. Diffusion echo-planar sequence was acquired with opposite phase encoding along the left-right axis, monopolar diffusion encoding with 128 diffusion-weighted images (2×64 for each encoding phase) at single shell maximum *b*=1500 s/mm^2^, 13 reference images at b=0 s/mm^2^, and 1.8mm isometric voxel size. See Supplementary Materials and Methods for detailed diffusion sequence and T1-weighted structural acquisition parameters. The preprocessing of the diffusion MRI data was computed according to well-established pipelines from the MRtrix3^32^ and FMRIB Software Library (FSL 6.0)^33^ toolboxes that have been described previously including in a recent publication from our laboratory^14^. Tensor-based quantitative maps of diffusion metrics (FA, MD, RD, and AD) were generated and used to perform Tract-Based Spatial Statistics (TBSS) whole-brain voxelwise analyses across all participants by creating a WM skeleton, which was registered to the Montreal Neurological Institute (MNI) standard space MNI152 template, from the averaged thresholded FA^34^.

### Statistical Analyses

Group comparisons for demographic, neuropsychological, and self-reported measures between CTL and iHUD were assessed with Student’s t-tests for continuous variables and Chi-square (χ^2^) tests for categorical variables (Table 1). Variables that reached the familywise error (FWE) threshold of p<.005 (.05/11 statistical tests) were considered to differ significantly between the groups. Commonly used drugs (alcohol and cannabis) were also compared between the groups using the same statistical tests with a FWE threshold of p<.008 (.05/6 statistical tests). Although nicotine use was assessed, no between-group comparisons were carried out for this measure because 33 iHUD and only one CTL participant reported being current smokers. Table 2 summarizes the treatment-related changes in behavioral measures as well as in both baseline and cue-induced drug craving variables. Paired t-tests with FWE-corrected thresholds were used to assess significant changes from MRI#1 to MRI#2 (p<.007 [.05/7 statistical tests] for the seven behavioral measures and p<.013 [.05/4 statistical tests] for the four drug craving variables). Comparisons reaching p<.05 were reported as trends for this analysis.

Whole-brain longitudinal WM analyses were carried out using the FSL tool “Randomise,” a general linear model for non-parametric permutation inferences^35^ using 10,000 permutations. To emphasize the investigation of the WM changes, a map of the between-scan “delta” (i.e., MRI#2 minus MRI#1; labeled as Δ) for each diffusion metric was computed and used in a design matrix coding for independent groups t-tests (two contrasts: CTL>HUD and CTL<HUD) while correcting for education, age, and intracranial volume (ICV), in line with common practices. ICV was estimated with the Freesurfer Software Suite (v7.2)^36^ from structural T1-weighted images. Since education differed between the groups and was correlated with WM diffusion metrics, this variable was added as a covariate in whole-brain analyses. To account for multiple comparisons, Threshold-Free Cluster Enhancement (TFCE) correction, which enhances areas of signal exhibiting spatial contiguity to better discriminate clusters, was applied.^37^ Neuroanatomical localization of WM tracts was performed with the FSL “atlasquery” toolbox and the “JHU ICBM-DTI-81 White-Matter Labels” atlas^38^, with an average probability of region overlap threshold of 2%.

### Other objectives

We tested whether the specific changes in WM diffusion metrics observed in the HUD group were associated with treatment-related outcomes including craving variables, selected behavioral measures, treatment adherence, and self-reported methadone/buprenorphine dosages. Targeted voxelwise correlations between the WM diffusion Δ metrics and the z-scored Δ values of the clinical outcome variables showing a significant change from MRI#1 to MRI#2 [i.e., baseline craving (pre-task) and cue-induced craving (picture/scene-induced)] were performed with a mask of the significant voxels where the HUD group showed increased FA at MRI#2 (i.e., 12,514 voxels). Although changes in picture-induced drug arousal and craving did not reach corrected significance levels and were therefore only considered trends, we included these measures in voxelwise correlation analyses in addition to therapy attendance, for completeness. The correlational analyses significance threshold was set to 1-p>.949 (TFCE-corrected). The magnitude of the correlations (r values) was estimated from the extracted WM metrics in the mask of significant voxels with IBM SPSS statistics version 27 (IBM Corp, Armonk, NY).

## Results

### Demographic, neuropsychological, and behavioral measures

The CTL and HUD groups did not differ significantly on age, gender, race, days between scans, and ICV, but did differ in education [CTL>HUD] (Table 1). Neuropsychological and self-reported measures highlighted the expected between-group differences in verbal IQ [CTL>HUD] as well as in depression and anxiety symptoms [HUD>CTL] (Table 1). The groups also differed on nicotine, alcohol, and cannabis use where more iHUD reported regular use of nicotine (not enough CTL were available for statistical comparison) and cannabis (inclusive of more years of regular cannabis use) with an opposite pattern for days of alcohol use in the past month (as expected from the controlled inpatient environment where the iHUD resided).

At MRI#2, the mean and SD of abstinence duration in iHUD were 249.3±231 days. There were significant decreases in both baseline craving and scene-induced craving intensity in the iHUD at MRI#2 compared to MRI#1 (Table 2). Trends for decreases with time were also observed for the drug picture-induced arousal and craving ratings. There were no significant longitudinal changes in the other select behavioral outcome measures (encompassing mood, sleep, and perceived stress) in the HUD group (Table 2). Independent longitudinal comparisons for positive and negative affect did not reach the corrected threshold (p=.054 and p=.066, respectively), but a time × affect ANOVA showed a significant interaction (p=.028) explained by trends for decreased negative and increased positive affect with treatment.

### Whole-brain white matter changes and correlations with clinical outcome measures

Figure 1 shows voxels where the Δ for the diffusion metrics were significantly different in iHUD than CTL. Specifically, compared to the CTL group, the HUD group showed a significant increase in FA (.949<1-p<.986) and decreases in MD (.949<1-p<.997) and RD (.949<1-p<.999) at MRI#2 as directly compared to MRI#1. No significant effects were found for AD. Increased FA in iHUD was localized to parts of the genu, body, and splenium of the corpus callosum as well as in bilateral anterior corona radiata, left superior corona radiata, right anterior limb of internal capsule, right posterior thalamic radiation, bilateral superior longitudinal fasciculus, right sagittal stratum, and right external capsule. Voxels showing decreased MD and RD in iHUD were more widespread in voxel count, although still found primarily in the same regions. Specific clustered results and localizations are summarized in Supplementary Table 1.

**Figure 1.**
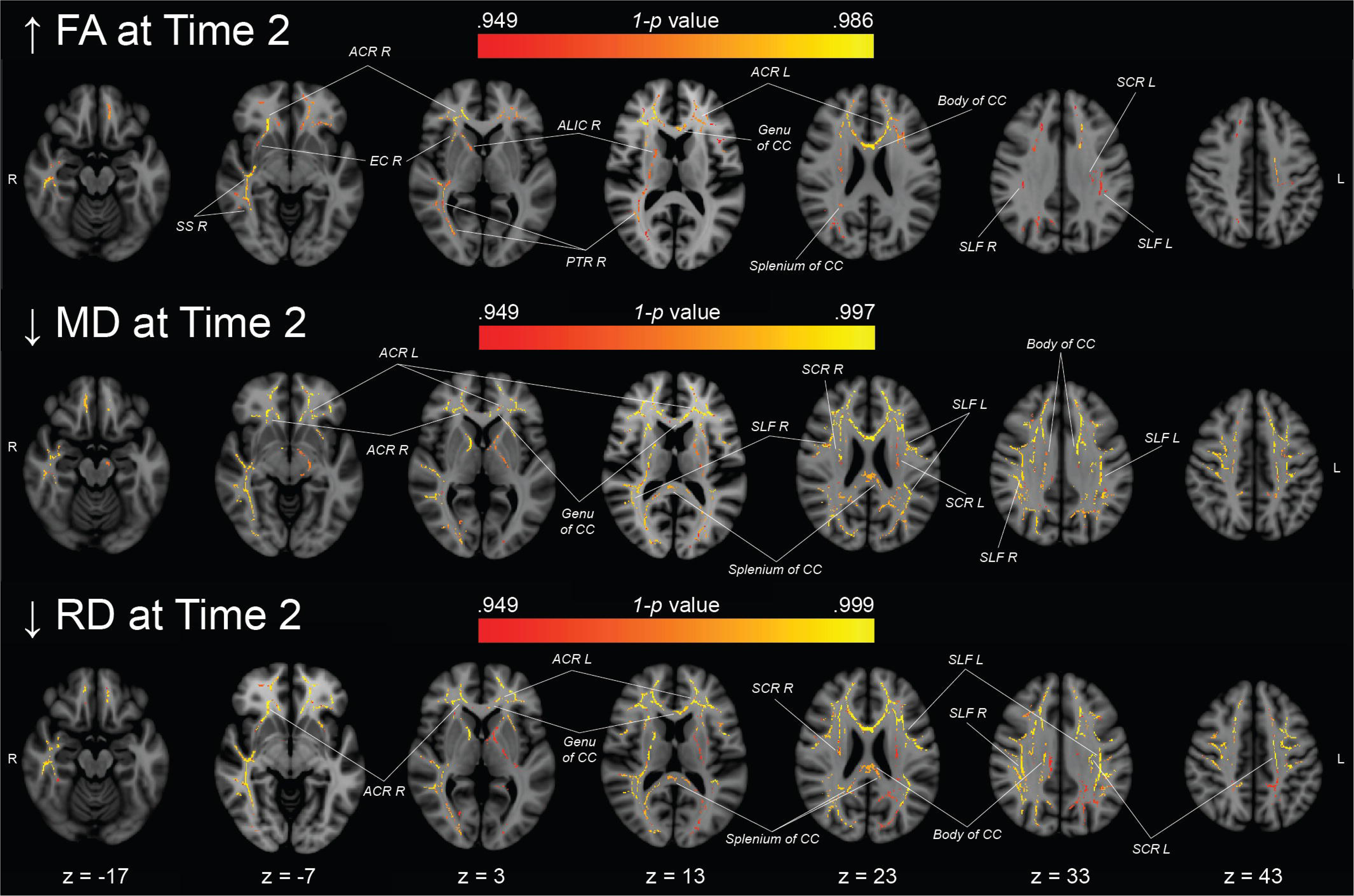
Group-specific whole-brain differences of fractional anisotropy (FA) and mean (MD) and radial (RD) diffusivities between two longitudinal MRI scans. Thresholded maps of significant voxels (1-p>.949, TFCE corrected) where the Δ between MRI#1 and MRI#2 differed between healthy controls and individuals with HUD. The first row shows significant regions where HUD had higher FA at MRI#2 vs. MRI#1 compared to CTL. The second and third rows show significant voxels where HUD showed decreases in MD and RD, respectively, at MRI#2 vs. MRI#1 as compared to CTL. See Supplementary Table 1 for clustered results. ACR: Anterior corona radiata, ALIC: Anterior limb of the internal capsule, CC: Corpus callosum, EC: External capsule, L: Left, PTR: Posterior thalamic radiation, R: Right, SCR: Superior corona radiata, SLF: Superior longitudinal fasciculus, SS: Sagittal stratum.

The between-scan Δ in FA, MD, and RD in the genu and body of the corpus callosum and the left anterior corona radiata were significantly correlated with the Δ of baseline craving (.949<1-p<.999) (Figure 2). Specifically, increased FA and decreased MD and RD at MRI#2 vs. MRI#1 were associated with decreased baseline craving. Correlations between the WM metrics and scene-induced craving Δ values did not yield significant results. Similarly, no correlations between the WM metrics Δ values with the other cue-induced craving measures, therapy attendance, and self-reported methadone/buprenorphine dosages reached significance (Supplementary Results).

**Figure 2.**
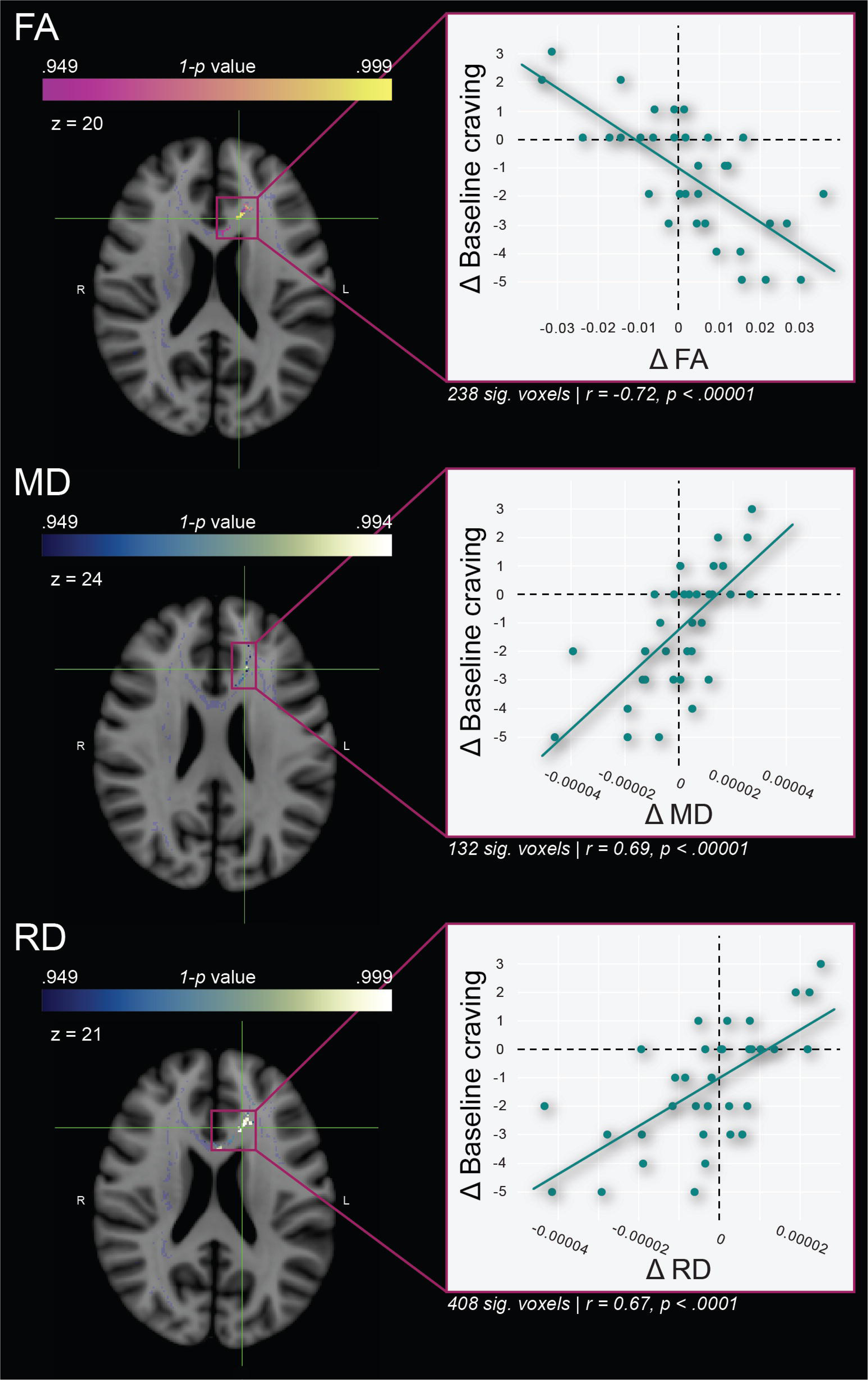
Voxelwise correlations between Δ in both the diffusion metrics and baseline craving in iHUD. Voxels where the increased FA and decreased MD and RD at MRI#2 significantly correlated with a reduction of baseline craving. This analysis used the mask of FA recovery showed in Figure 1 (grey). These effects were observed mainly in the genu and body of the corpus callosum and the left anterior corona radiata. Green crosshairs indicate the peak voxel location for the correlation with each of the three metrics (MNI coordinates, FA: x=-13, y=23, z=20; MD: x=-16, y=26, z=24; RD: x=-15, y=24, z=21).

## Discussion

The goal of this study was to longitudinally investigate the WM changes with inpatient treatment in iHUD undergoing MAT, as well as changes in baseline and cue-induced craving measures and behavioral clinical measures inclusive of mood/affect, sleep, and stress. Our main findings demonstrate that neuroanatomical impairments in distributed frontal WM tracks in iHUD undergo appreciable change characterized by increased FA and decreased MD and RD, which are indicative of recovery processes. Our findings also show that recovery of WM microstructure, specifically in the genu and body of the corpus callosum and bilateral anterior corona radiata, correlated with reductions in baseline drug craving.

Multiple studies show WM impairments across various drugs of abuse including heroin^15–19,21^, cocaine^39–42^, alcohol^43,44^, nicotine^45–47^, and cannabis^48^. Cross-sectional studies have suggested a correlation between abstinence and potential WM recovery, including our prior study in cocaine use disorder in which, compared to active users, abstainers (representing a group with longer abstinence durations and/or less chronicity of use) showed limited impairments when compared to CTL^14^. Corroborating results reported less WM impairments in long-term drug abstainers (negative urine status for any drugs) compared to iHUD undergoing methadone maintenance treatment^49^, and in heroin abstainers compared to individuals who relapsed to heroin use^17^.

Although limited in number, longitudinal studies corroborate these cross-sectional studies^21^. Tractography studies using atlas-based analyses showed increased FA values in the fronto-striatal circuits and nucleus accumbens fiber tracts after eight versus two months of abstinence in heroin users (without MAT)^22,23^. The current study bolstered the FA increase in these networks during recovery, extending results to other diffusion metrics (MD and RD), in addition to showing a more widespread pattern than previously observed. These broad WM changes could stem from the relatively enriched services provided to these iHUD (all enrolled in an intensive inpatient biopsychosocial support program that included MAT). Taken together, this pattern is consistent with normalization/regeneration of axonal processes and remyelination^50–55^ in specific WM tracts with inpatient MAT in iHUD. As potential mechanisms, of mention are the chronic opiate effects (and their change with treatment) on neuroinflammation as mediated by microglia^56,57^ and gene expression in oligodendrocytes, the glia responsible for myelinating axons in the brain^58–62^.

The clinical relevance of these results also derives from the significant association of the longitudinal WM changes with parallel decreases in baseline craving. Based on the localization of the significant voxels, encompassing potential corticothalamic/corticolimbic circuits in the left anterior corona radiata and parts of the genu and body of the corpus callosum, these WM changes could relate to the recovery of higher-order cognitive functions including emotional regulation and top-down executive control^63,64^. Our findings suggest that more efficient communication in these areas could contribute to reduction in baseline, though not stimulus/cue-induced, craving^65^.

Our results should be interpreted in light of several limitations. First, replication in a larger independent sample is necessary as participants in this study were also included in previous cross-sectional reports^14,66^. Larger studies are also needed to examine effects of individual differences related to sex/gender, treatment-seeking status, and medication (e.g., methadone or buprenorphine) during recovery on WM microstructure. Although we found no significant relationship between baseline self-reported methadone/buprenorphine dosages and our dependent measures, more rigorous assessments of recovery effects related to MAT type and dosage should be implemented in future studies. Finally, the model on which TBSS analyses rely is known to lack specificity in areas with complex WM fiber architecture (including crossing fibers)^67^, precluding the observation of potentially relevant circuitry especially in subcortical WM regions.

This is the first study to investigate longitudinal whole-brain WM changes with MAT in iHUD. This study demonstrated increased FA, and decreased MD and RD, in fronto-striatal WM tracts after approximately 15 weeks in MAT in iHUD, consistent with normalization/regeneration of WM microstructure. This study also revealed an association between the time-dependent changes in the WM metrics with changes in baseline measures of craving. The association of WM recovery with the decrease of baseline craving at follow-up further suggests cognitive improvements with abstinence/treatment in iHUD, which may contribute to longer-term abstinence and relapse prevention.

## Supporting information

Supplementary Methods and Results

## Data Availability

All data produced in the present study are available upon reasonable request to the authors

## Author Contributions

The corresponding author, Rita Z. Goldstein, had full access the data in the study and takes responsibility for the integrity of the data and the accuracy of the data analysis.

*Concept and design:* Pierre-Olivier Gaudreault, Ahmet O. Ceceli, Nelly Alia-Klein, Rita Z. Goldstein

*Acquisition, analysis, or interpretation of data:* Ahmet O. Ceceli, Yuefeng Huang, Greg Kronberg, Pierre-Olivier Gaudreault, Sarah G. King, Nelly Alia-Klein, Rita Z. Goldstein

*Drafting of the manuscript:* Pierre-Olivier Gaudreault, Rita Z. Goldstein

*Critical revision of the manuscript for important intellectual content:* Sarah G. King, Yuefeng Huang, Ahmet O. Ceceli, Greg Kronberg, Nelly Alia-Klein, Rita Z. Goldstein

*Statistical analysis:* Pierre-Olivier Gaudreault

*Administrative, technical, or material support:* Ahmet O. Ceceli, Pierre-Olivier Gaudreault, Sarah G. King, Yuefeng Huang, Greg Kronberg, Nelly Alia-Klein, Rita Z. Goldstein

*Supervision:* Nelly Alia-Klein, Rita Z. Goldstein

