## Supplementary Methods and Results for "FRONTAL WHITE MATTER CHANGES INDICATE RECOVERY WITH INPATIENT TREATMENT IN HEROIN ADDICTION"

*These authors contributed equally.

**SUPPLEMENTARY MATERIAL**

**Number of figures**: 0

**Number of tables**: 1

**Corresponding author:**

Rita Z. Goldstein, PhD

Mount Sinai Professor in Neuroimaging of Addiction

Department of Psychiatry (primary) and Department of Neuroscience (secondary)

Chief, Neuropsychoimaging of Addiction and Related Conditions (NARC) Research Program

Icahn School of Medicine

Gustave L. Levy Place, Box 1230

New York, NY 10029

**Supplementary Materials and Methods**

***Exclusion Criteria***

Exclusion criteria were: 1) present or past history of DSM-5 diagnosis of psychotic disorder or neurodevelopmental disorder; 2) history of head trauma with loss of consciousness (>30 min); 3) history of neurological disorders including seizures; 4) current medical illness and/or evident infection including cardiovascular disease (e.g., high blood pressure), as well as metabolic, endocrinological, oncological, or autoimmune diseases, and infectious diseases common in individuals with substance use disorder including Hepatitis B and C or HIV/AIDS; 5) MRI contraindications including any metallic implants, pacemaker device, or pregnancy; 6) Individuals undergoing court-mandated treatment; 7) MRI quality assurance, including the presence of incidental findings in the WM as indicated by a radiologist, bad diffusion data, or an MRI session that did not include a diffusion sequence (1 HUD). We did not exclude individuals with HUD for history of other substance use disorder (e.g., alcohol, marijuana, stimulants/other opiates) or other psychiatric disorders with high rates of comorbidity with drug addiction (e.g., depression, post-traumatic stress disorder). The same exclusion criteria applied to the CTL except any history of substance use disorder was prohibitive.

***Comorbidities in individuals with heroin use disorder***

Comorbidities in iHUD included cocaine use disorder (n=4), sedative use disorder (n=4), alcohol use disorder (n=3), cannabis use disorder (n=2), meth/amphetamine use disorder (n=1), polysubstance use disorder (i.e., dependence on at least three groups of substances, not including nicotine and caffeine, in the past 12 months, with no single substance predominating; n=1), major depressive disorder (n=2), post-traumatic stress disorder (n=2), binge eating disorder (n=1), and panic disorder (n=1).

***Treatment-specific details***

All iHUD were undergoing medically-assisted treatment in an inpatient program at the time of the study. As a part of the clinical trial (NCT04112186), participants were also randomized to one of two 8-week treatment groups in addition to their regular treatment regimen. One group involved a mindfulness-based self-awareness and emotion regulation training (e.g., cognitive reappraisal of negative and savoring of positive thoughts/contexts); these strategies were delivered with the goal of diminishing drug cue-reactivity while enhancing natural reward processing and cognitive control over craving and drug-seeking behaviors (therapy details^1^). The other group involved a support training that included structured, therapist-guided and addiction-related psychoeducation, emotional expression, and discussions. Both groups also received daily 15-min practice sessions, guided by audio instructions or independent journaling exercises about addiction-related topics.

***Neuropsychological and Self-reported measures***

Neuropsychological and self-reported measures included estimated verbal [reading subtest of the Wide Range Achievement Test-3^2^] and non-verbal IQ [Matrix Reasoning subtest of the Wechsler Abbreviated Scale of Intelligence^3^] and handedness [modified Edinburgh Handedness Inventory^4^].

***Repeated behavioral measures***

Depression symptoms were assessed at both timepoints with the Beck Depression Inventory (BDI)^5^ and anxiety symptoms with the Beck Anxiety Inventory (BAI).^6^ Positive and negative affect were measured with the Positive and Negative Affect Schedule,^7^ and the Perceived Stress Scale^8^ assessed general stress levels in the past month.

***MRI Acquisition parameters***

MRI acquisitions were performed at both timepoint using the same Siemens 3.0 Tesla Skyra scanner (Siemens Healthineers AG, Erlangen, Germany) with a 32-channel head coil. Diffusion echo-planar sequence was acquired with opposite phase encoding along the left-right axis, monopolar diffusion encoding with 128 diffusion-weighted images (2×64 for each encoding phase) at single shell maximum *b*=1500 s/mm^2^, 13 reference images at b=0 s/mm^2^, field of view (FOV)=882×1044 mm, , repetition time (TR)=3650ms, echo time (TE)=87ms, bandwidth=1485 Hz/px, and 80° flip angle, multiband=3, no in-plane acceleration. Structural T1-weighted images were acquired using an MPRAGE sequence [sagittal orientation, FOV=256×256×179mm^3^; 0.8mm isotropic resolution; TR=2400ms; TE=2.07ms; inversion time 1000ms; flip angle 8° with binomial (1, -1) fat saturation; bandwidth 240 Hz/pixel; 7.6ms echo spacing, and in-plane acceleration (GRAPPA) factor of 2].

**Supplementary Results**

***Correlations with therapy and drug craving measures that did not reach statistical significance***

Although only trends were observed for their changes from MRI#2 – MRI#1 (Δ), we performed voxelwise correlations between the picture-induced drug arousal and craving ratings of drug cues and the Δ WM metrics with no significant correlations. There was a trend (1-p>0.9) for a correlation between the Δ RD and picture-induced drug arousal ratings (1-p<.941) and between the Δ MD and picture-induced drug craving ratings (1-p<.932) such that the lower the Δ diffusivity metrics the lower the delta drug arousal/craving ratings. No other significant effects or trends were observed for picture-induced arousal [(FA: 1-p<.896, MD: 1-p<.848, AD: 1-p<.516) or craving (FA: 1-p<.673, AD: 1-p<.845, RD: 1-p<.875), respectively]. A trend was also observed for the correlation between the Δ FA and methadone/buprenorphine dosage (1-p<.937) such that the lower the Δ FA the higher the dosage, with no other significant effects or trends (MD: 1-p<.406, AD: 1-p<.653, RD: 1-p<.706). No significant correlations were found between the Δ WM metrics and therapy attendance (FA: 1-p<.723, MD: 1-p<.377, AD: 1-p<.404, RD: 1-p<.488).

***Supplementary Table 1*** *– Clustered between-group white matter differences on Δ diffusion metrics between MRI#1 and MRI#2.*

|  |  |  |  | Commissural | | | | Projection | | | | | | | | Association | | | | | | | | Brainstem | | | | |
| --- | --- | --- | --- | --- | --- | --- | --- | --- | --- | --- | --- | --- | --- | --- | --- | --- | --- | --- | --- | --- | --- | --- | --- | --- | --- | --- | --- | --- |
|  | Effect | Voxels | Max 1-*p* | GCC | BCC | SCC | TAP | ACR | SCR | PCR | ALIC | PLIC | RLIC | PTR | CP | SLF | SFO | UF | SS | EC | CgC | CgH | FX / ST | CST | ML | ICP | MCP | SCP |
| Fractional Anisotropy | |  |  |  |  |  |  |  |  |  |  |  |  |  |  |  |  |  |  |  |  |  |  |  |  |  |  |  |
| Cluster 1 | ΔHUD > ΔCTL | 11,807 | .986 | **X** | **X** |  |  | **B** |  |  | R |  |  | R |  |  |  |  | R | R |  |  |  |  |  |  |  |  |
| Cluster 2 | ΔHUD > ΔCTL | 595 | .956 |  |  |  |  |  | **L** |  |  |  |  |  |  | **L** |  |  |  |  |  |  |  |  |  |  |  |  |
| Cluster 3 | ΔHUD > ΔCTL | 75 | .955 |  |  |  |  |  |  |  |  |  |  |  |  | **R** |  |  |  |  |  |  |  |  |  |  |  |  |
| Cluster 4 | ΔHUD > ΔCTL | 43 | .953 |  |  | **X** |  |  |  |  |  |  |  |  |  |  |  |  |  |  |  |  |  |  |  |  |  |  |
| Mean Diffusivity | |  |  |  |  |  |  |  |  |  |  |  |  |  |  |  |  |  |  |  |  |  |  |  |  |  |  |  |
| Cluster 1 | ΔCTL > ΔHUD | 23,557 | .997 | X | X | X |  | B | B |  |  |  |  |  |  | B |  |  |  |  |  |  |  |  |  |  |  |  |
| Radial Diffusivity | |  |  |  |  |  |  |  |  |  |  |  |  |  |  |  |  |  |  |  |  |  |  |  |  |  |  |  |
| Cluster 1 | ΔCTL > ΔHUD | 32,668 | .999 | X | X | X |  | B | B |  |  |  |  |  |  | B |  |  |  |  |  |  |  |  |  |  |  |  |

**Notes**: Significance for group differences was considered at 1-p > .949 with a clustering minimum of 30 voxels. The right part of the table represents the averaged probabilities of all significant voxels from a cluster to overlap with the “JHU ICBM-DTI-81 White-Matter Atlas” labels. Specific regions are divided into types of fibers (commissural, projection, association, and tracts in the brainstem). Localization of significant regions was identified with an overlap threshold of 2%. “X” is used for non-lateralized regions. “L/R/B” correspond to Left / Right / Bilateral regions. Bold and underline cases represent overlap probabilities >5%.

**Acronyms (in the presented order)**: [Commissural fibers] GCC: genu of corpus callosum, BCC: body of corpus callosum, SCC: splenium of corpus callosum, TAP: tapetum. [Projection fibers] ACR: anterior corona radiata, SCR: superior corona radiata, PCR: posterior corona radiata, ALIC: anterior limb of internal capsule, PLIC: posterior limb of internal capsule, RLIC: retrolenticular part of internal capsule, PTR: posterior thalamic radiation, CP: cerebral peduncle. [Association fibers] SLF: superior longitudinal fasciculus, SFO: superior fronto-occipital fasciculus, UF: uncinate fasciculus, SS: sagittal stratum, EC: external capsule, CgC: cingulum in the cingulate cortex, CgH: cingulum in the hippocampal region, FX / ST: fornix/stria terminalis. [Tracts in the brainstem] CST: corticospinal tracts, ML: medial lemniscus, ICP: inferior cerebellar peduncle, MCP: middle cerebellar peduncle, SCP: superior cerebellar peduncle.
